# Decreased door-to-balloon time in patients with ST-segment elevation myocardial infarction during the early COVID-19 pandemic in South Korea – an observational study

**DOI:** 10.1101/2021.09.17.21263760

**Authors:** Sukhyun Ryu, Dasom Kim, Lae Young Jung, Baekjin Kim, Chang-Seop Lee

## Abstract

**Background:** The coronavirus disease 2019 (COVID-19) resulted in a marked decrease in the number of patient visits for acute myocardial infarction (AMI) and delayed patient response and intervention in several countries. This study evaluated the effect of the COVID-19 pandemic on the number of patients, patient response time (pain-to-door), and intervention time (door-to-balloon) for patients with ST-segment elevation myocardial infarction (STEMI) and non-ST-segment elevation myocardial infarction (NSTEMI).

**Methods:** Patients with STEMI or NSTEMI visiting a hospital in South Korea who underwent primary coronary intervention during the COVID-19 pandemic (January 29, 2020, to December 31, 2020) were compared with those in the equivalent period in 2018 to 2019. Patient response and intervention times were compared for the COVID-19 pandemic window (2020) and the equivalent period in 2018 to 2019.

**Results:** We observed no decrease in the number of patients with STEMI (*P*=0.50) and NSTEMI (*P*=0.94) during the COVID-19 pandemic compared to that in the previous years. Patient response times (STEMI: *P*=0.34; NSTEMI: *P*=0.89) during the overall COVID-19 pandemic period did not differ significantly. However, we identified a significant decrease in time to intervention among patients with STEMI (14%; p<0.01) during the early COVID-19 pandemic.

**Conclusions:** We found that the number of patient with STEMI and NSTEMI was consistent during the COVID-19 pandemic and that no time delays in patient response and intervention occurred. However, the door-to-balloon time among patients with STEMI significantly reduced during the early COVID-19 pandemic, which could be attributed to reduced emergency care utilization during the early pandemic.

## INTRODUCTION

Healthcare resources have largely been focused towards response to the coronavirus disease 2019 (COVID-19) pandemic, and concerns regarding decreased availability of medical care for patients with acute myocardial infarction (AMI) have emerged globally.^1-3^ Considering the possibility of a future pandemic or emergent health scenarios and that healthcare utilization and healthcare systems for acute medical conditions vary by country, identifying the effect of the COVID-19 pandemic on patient response and treatment for acute medical conditions such as AMI is crucial.^4^

AMI can be divided into subgroups of ST-segment elevation myocardial infarction (STEMI) and non-ST-segment elevation myocardial infarction (NSTEMI). STEMI is an acute medical condition commonly caused by thrombotic occlusion of the coronary artery, and it is a fatal cardiovascular emergency that requires rapid percutaneous coronary intervention (PCI) within 90 minutes.^5^ NSTEMI manifests varying symptoms on presentation and disparate clinical outcomes depending on whether the intervention is early or delayed; PCI has been recommended within 72 hours, depending on the patient’s risk category.^6^

In many countries, the time taken from pain onset to first medical contact (i.e., pain-to-door time) among patients with STEMI and in-hospital delivery of revascularization (i.e., door-to-balloon time at the coronary intervention laboratory) during the early COVID-19 pandemic differed drastically compared to the same period in the previous year^7-9^.

Here, to determine the effect of the COVID-19 pandemic on patient response and intervention for AMI, we examined the time to patient response and PCI among patients with STEMI and NSTEMI on 3 different epidemic waves of COVID-19 in South Korea.

## METHODS

We conducted a single-centre, retrospective observational study including patients with STEMI and NSTEMI visiting the Jeonbuk National University hospital between January 1, 2018, to December 31, 2020. The hospital has a tertiary care cardiovascular centre that offers 24/7 coronary intervention covering the North Jeolla province, which has a population of 2 million people. Patients with AMI admitted through the emergency department of the hospital were included in this study. Patients were identified using ICD-10 codes for STEMI (I21.0, I21.1, I21.2, and I21.3), NSTEMI (I21.4 and I22.2), and unspecified (I21.9).

We used the pain-to-door time to assess the time to patient response. Pain-to-door time was identified as the interval from the onset of AMI-related symptoms, including chest discomfort, to the time of first medical contact. Furthermore, we used the door-to-balloon time to examine the time to PCI. The door-to-balloon time was defined as the interval from arrival at the emergency department to successful wire crossing of the culprit lesion through PCI. These proxies are commonly used to assess the patient’s health-seeking behavior and to assess emergency cardiac care, respectively.^10^

To determine the temporal changes in pain-to-door and door-to-balloon times, we divided the study duration into 3 periods (Period-1: epidemiologic weeks 4–19, Period-2: 20– 33, and Period-3: 34–52) basis the official COVID-19 outbreak characteristics in South Korea.^11 12^ The incidence rate ratio with 95% confidence intervals (CIs) of patients with STEMI and NSTEMI across the 3 periods was estimated using the weekly count of patients with STEMI and NSTEMI between January 2018 and December 2020. The pain-to-door and door-to-balloon times were tested for normality and presented as medians with interquartile ranges (IQRs). To evaluate the difference in the number of patients with STEMI and NSTEMI and in patient demographics between pre-COVID-19 (2018 and 2019) and COVID-19 pandemic (2020) periods, we conducted an analysis of variance (ANOVA), Kruskal– Wallis one-way ANOVA or chi-square test, as appropriate. Furthermore, to determine the difference in the times between pre-COVID-19 and COVID-19 pandemic periods, we conducted a Welch 2-sample t-test or Mann-Whitney test, as appropriate. P<0.05 was considered statistically significant, and all analyses were performed using R version 3.6.1 (R Foundation for Statistical Computing, Vienna, Austria).

## RESULTS

In total, 831 patients with acute myocardial infarction visited the cardiovascular centre via the emergency department between 2018 and 2020. We excluded 23 patients who did not have indications for STEMI or NSTEMI. A total of 808 patients were included, of which 439 (54.3%) experienced STEMI and 369 (45.7%) experienced NSTEMI.

We did not observe any significant differences in patient demographics, comorbidities, and history of myocardial infarction between patients with STEMI and NSTEMI (Table 1). No significant difference in the numbers of patients with STEMI and NSTEMI during the COVID-19 pandemic in 2020 and the identical periods in years 2018 and 2019 were observed (2018, P=0.50; 2019, P=0.94; Table 1; Figure 1). Furthermore, during the 3 different epidemic periods of COVID-19 in South Korea (Figure 2A), we did not observe a significant difference in the incidence rate ratio of STEMI (Period-1: 1.1, 95% CI 0.7–1.8; Period-2: 1.1, 95% CI 0.7–1.7; Period-3: 0.9, 95% CI 0.6–1.3) and NSTEMI (Period-1: 0.9, 95% CI 0.5–1.5; Period-2: 1.2, 95% CI 0.7–2.1; Period-3: 0.9, 95% CI 0.6–1.3) (Figure 2B).

**Table 1.** Demographic characteristics of patients with ST-segment elevation myocardial infarction (STEMI) and non-ST-segment elevation myocardial infarction (NSTEMI) from 2018 to 2020.

|  | STEMI |  |  |  | NSTEMI |  |  |  |
| --- | --- | --- | --- | --- | --- | --- | --- | --- |
| Variable | 2018 | 2019 | 2020 | *P* value | 2018 | 2019 | 2020 | *P* value |
| No. (%) | 138<br>(31.44) | 155<br>(35.31) | 146<br>(33.26) | 0.50 | 124<br>(33.60) | 125<br>(33.88) | 120<br>(32.52) | 0.94 |
| Demographics |  |  |  |  |  |  |  |  |
| Age, mean (SD), y | 63.96<br>(13.77) | 67.19<br>(11.69) | 65.36<br>(12.57) | 0.09 | 67.55<br>(11.84) | 68.52<br>(12.35) | 65.08<br>(12.50) | 0.08 |
| Male sex | 106 | 125 | 119 | 0.58 | 87 | 81 | 88 | 0.91 |
| Obesity (high BMI) | 6 | 7 | 5 | 0.88 | 5 | 6 | 0 | 0.11 |
| Comorbidities |  |  |  |  |  |  |  |  |
| Hypertension | 72 | 82 | 76 | 0.99 | 77 | 66 | 60 | 0.14 |
| Diabetes | 230 | 45 | 38 | 0.43 | 52 | 44 | 50 | 0.47 |
| Smoker | 83 | 95 | 89 | 0.93 | 51 | 63 | 60 | 0.22 |
| Prior MI | 7 | 16 | 11 | 0.39 | 17 | 25 | 27 | 0.19 |

STEMI, ST-segment elevation myocardial infarction; NSTEMI, non-ST-segment elevation myocardial infarction; SD, standard deviation; BMI, body mass index; MI, myocardial infarction.

**Figure 1.**
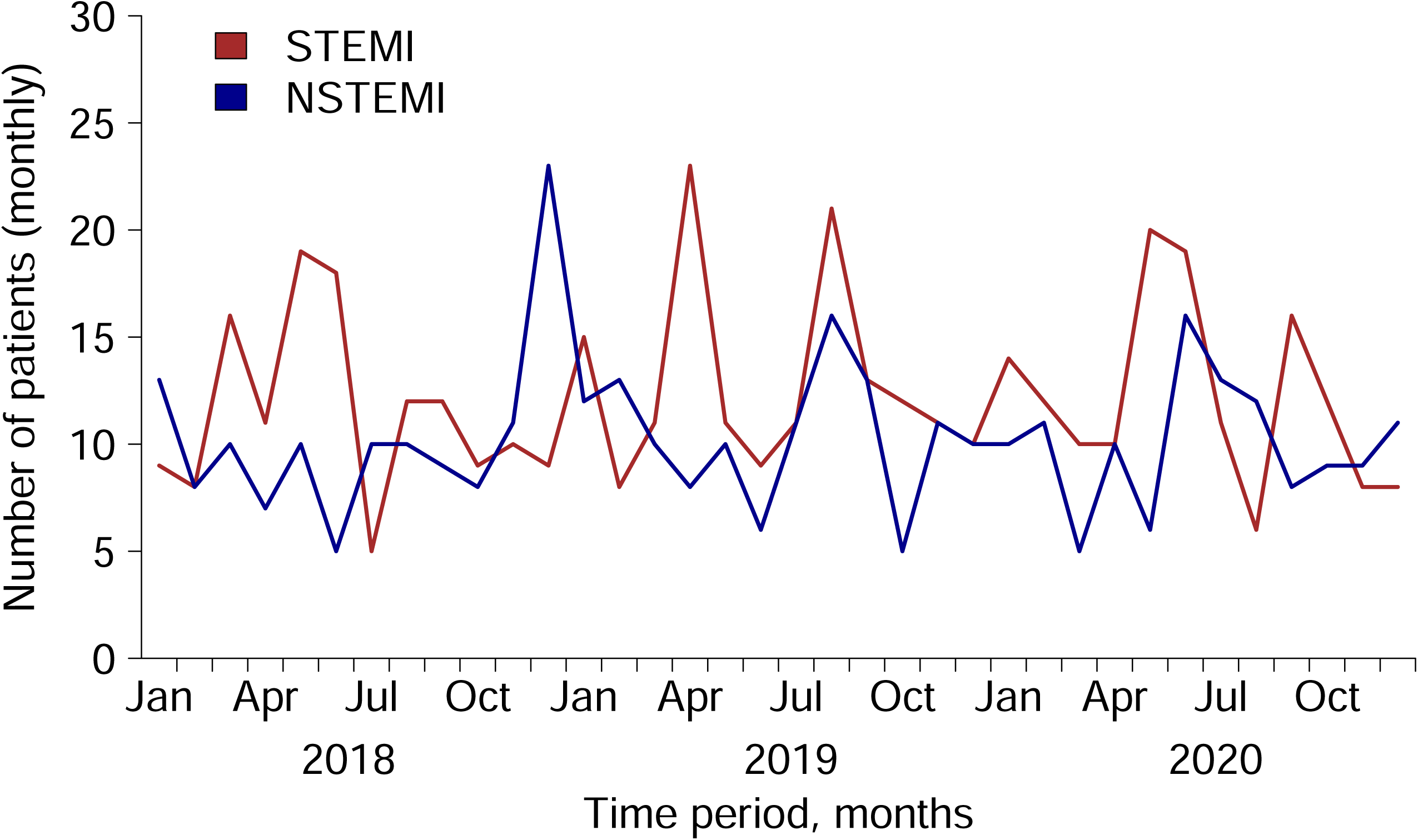
The monthly count of patients with ST-segment elevation myocardial infarction (STEMI) and non-ST-segment elevation myocardial infarction (NSTEMI) from 2018 to 2020. STEMI = ST-segment elevation myocardial infarction, NSTEMI = non-ST-segment elevation myocardial infarction.

**Figure 2.**
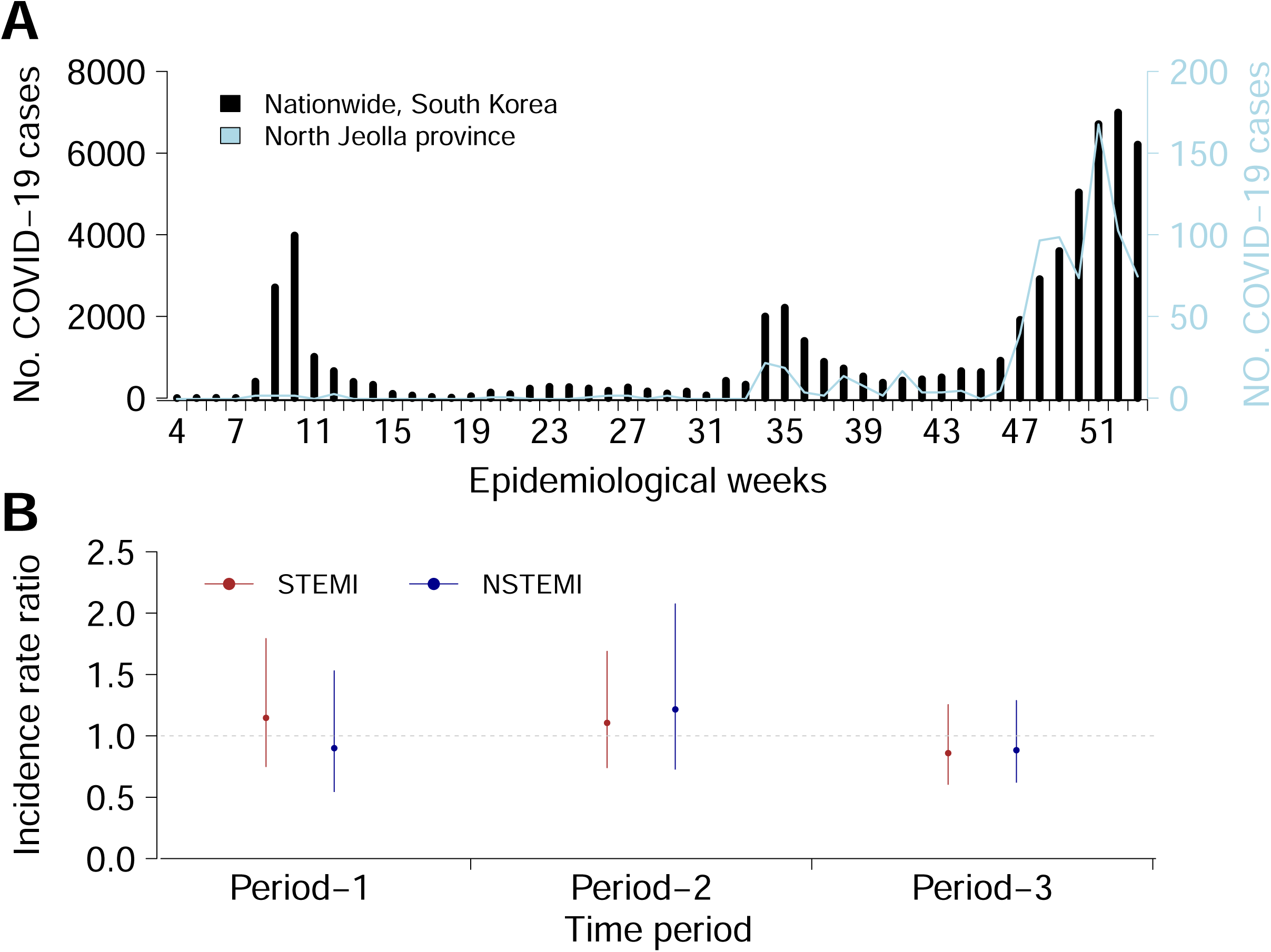
Coronavirus 2019 (COVID-19) cases in South Korea and 2018/2019-2020 estimated rate ratio of hospitalised patients with ST-segment elevation myocardial infarction (STEMI) and non-ST-segment elevation myocardial infarction (NSTEMI). (A) The weekly count of local COVID-19 cases nationwide (black bar) and in the North Jeolla province (solid blue line). (B) The 2018/2019-2020 estimated admission rate ratio of patients with STEMI (brown bar) and NSTEMI (blue bar) during the 3 different epidemic periods of COVID-19. The bar indicates 95% confidence intervals. The study period included Period-1 (epidemiological week 4-19), Period-2 (week 20-33), and Period-3 (week 34-52). STEMI = ST-segment elevation myocardial infarction. NSTEMI = non-ST-segment elevation myocardial infarction.

For patients with STEMI, the median pain-to-door time was 120 min (IQR: 65-235 min) during the COVID-19 pandemic in 2020 and 125 min (IQR: 67-232 min) from 2018 to 2019. We did not observe a statistical difference in the pain-to-door time during the 3 periods of the COVID-19 pandemic (Period-1: P=0.80, Period-2: P=0.73, and Period-3: P=0.56; Figure 3A). However, we noted that the door-to-balloon time significantly decreased during Period-1 in 2020 (median, 37 min; IQR 32-45 min; P=0.004) than in 2018 or 2019 (median, 43 min; IQR 36-64 min). We observed no difference in the door-to-balloon time in Period-2 (P=0.17) and Period-3 (P=0.94) (Figure 3B).

**Figure 3.**
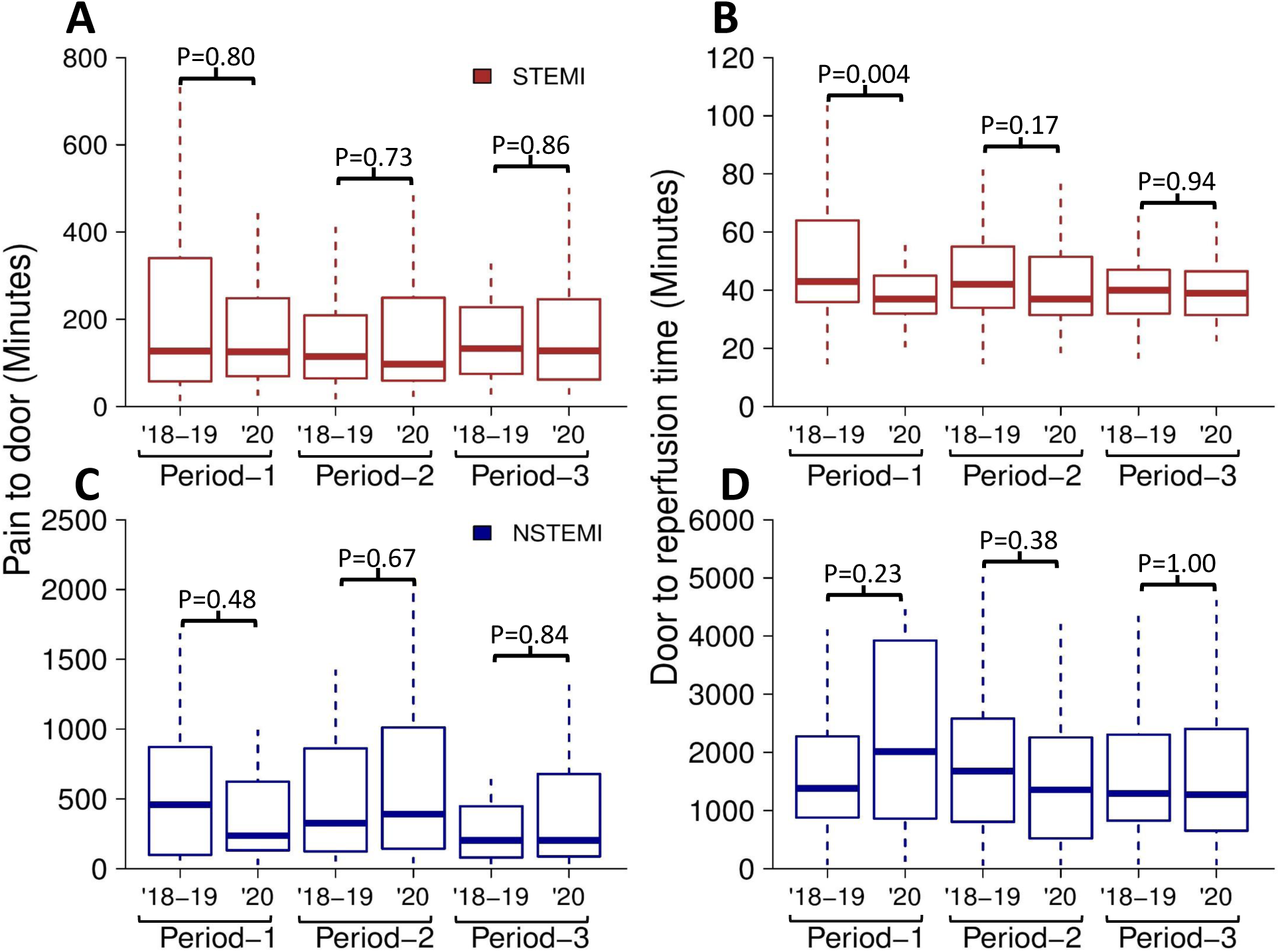
Bar plot of the pain-to-door and door-to-reperfusion times of patients with ST-segment elevation myocardial infarction (STEMI) and non-ST-segment elevation myocardial infarction (NSTEMI) during 3 different periods in South Korea. The study period included Period-1 (epidemiological week of 4-19), Period-2 (week of 20-33), and Period-3 (week of 34-52). STEMI = ST-segment elevation myocardial infarction. NSTEMI = non-ST-segment elevation myocardial infarction.

For patients with NSTEMI, the median pain-to-door time was 265 min (IQR, 103-630 min) during the COVID-19 pandemic and 290 min (IQR, 105-708 min) for the same periods in 2018 and 2019. We did not observe a significant difference in pain-to-door times across the epidemic periods of COVID-19 (Period-1: P=0.48, Period-2: P=0.67, and Period-3: P=0.84; Figure 3C). The median door-to-ballon time for patients with NSTEMI was 1,388 min (IQR, 675-2647 min) during the COVID-19 pandemic and 1,375 min (IQR, 823-2379 mins) for the identical periods in 2018 and 2019. We did not observe a significant difference in door-to-balloon times during Period-1 (P=0.23), Period-2 (P=0.38), and Period-3 (P=1.00) (Figure 3D).

## DISCUSSION

A significant decrease in the number of patients with AMI was observed in many countries during the early COVID-19 pandemic.^13^ This observation could be attributed to patient anxiety and desire to avoid contact with patients with COVID-19 in the medical facility during the COVID-19 pandemic, which likely affected the patients’ care-seeking behavior.^14^ Second, a lockdown strategy to enforce staying at home in these countries likely discouraged patients from seeking medical care.^14 15^ Third, a substantially decreased incidence of viral respiratory infections, including influenza,^16^ might have decreased the incidence of AMI in these countries.^17 18^

Unlike previous studies, we observed no significant decrease in the number of patients with STEMI and NSTEMI. Furthermore, we observed no significant delay in patient response time across the 3 different epidemic periods of COVID-19 in our study. Demographic characteristics and patient comorbidities are known to contribute to AMI.^15^ However, in the present study, we did not identify differences in demographic characterisitcs and comorbidities of patients between the COVID-19 pandemic period and those in the previous years. In South Korea, a lockdown strategy was not implemented for the overall COVID-19 pandemic period.^19 20^ Our finding is consistent with that of a previous study, which reported that countries that implemented partial lockdowns against the COVID-19 pandemic did not observe an effect in the number of patients with STEMI.^13^ Therefore, this finding suggests that the COVID-19 pandemic had a limited effect on the healthcare-seeking behavior of patients with STEMI and NSTEMI. Our finding is consistent with that of a previous report that the number of emergency department visits by patients with severe medical conditions did not reduce during the COVID-19 pandemic in South Korea.^21^

The results of this study indicate that patient response time did not differ significantly across the 3 different epidemic periods of the COVID-19 pandemic, and this finding supports the hypothesis that the COVID-19 pandemic had a limited effect on the healthcare-seeking behavior of patients with STEMI and NSTEMI.

We observed that the door-to-balloon time significantly reduced (14%) during the eaely COVID-19 pandemic (Period-1). Previous Korean studies reported decreased number of patient-visit to emergency department (46%-77%) during the early COVID-19 pandemic. ^21-23^ Therefore, the reduction of the door-to-balloon time could be attributed to decreased emergency care utilisation during the early COVID-19 pandemic as patients with low severity defer emergency department visits owing to the fear of the infection in the medical facility.^22 23^ This decrease in emergency care utilisation was also observed during the MERS outbreak in 2015 in South Korea.^24^

We observed that the pain-to-door and door-to-balloon times among patients with NSTEMI varied in a greater range than for those among patients with STEMI. Patients with NSTEMI typically defer the intervention until other underlying conditions are managed or staff hours are available.^6^ This could have affected the results of our study, possibly resulting in greater variation in door-to-ballonn time among patients with NSTEMI. However, we observed no significant difference between pre-COVID-19 and the COVID-19 pandemic years.

This study had limitations. First, we considered the number of patients in our study as a proxy of the incidence of AMI in the North Jeolla Province. Second, we did not account for patients that transferred to regions outside of the North Jeolla province. However, our study also had several strengths. First, this is the first study to assess changes in the number of patients with STEMI and NSTEMI in South Korea, where no lockdown strategy was incorporated as a measure against the COVID-19 pandemic. Furthermore, our study measured the time between symptom onset to first medical contact and first medical contact to PCI, which is the first evaluation of this kind during the specified time period for patients with STEMI and NSTEMI in South Korea.

## CONCLUSION

Our study shows that the number of patients with AMI did not decrease and patient’s response and intervention times did not increase. However, the time to intervention since first medical contact decreased significantly among patients with STEMI during the early COVID-19 pandemic. Further study is needed to identify the factors contributing to this decrease.

## Data Availability

The datasets used in the current study are available at https://github.com/gentryu/Korean_AMI_COVID19.

## Acknowledgments

The authors thank Mir Jeon for her kind assistance in collecting the data during the current study.

## Contributors

SR: research design, editing article from an epidemiological perspective and authorship of the first and final draft. DK, BK: statical analysis and statistical analysis. LYJ: data collection and editing the article from an acute coronary syndrome perspective. CSL: senior author who proposed the research question, research design, assisted in authorship of the manuscript.

## Patient consent for publication

Not required.

## Ethics approval

This study was approved by the Institutional Review Board at the Jeonbuk National University Hospital (IRB No. 2021-03-043).

## Provenance and peer review

Not commissioned; externally peer reviewed

